# Comparison of caffeine consumption behavior with plasma caffeine levels as exposures in drug-target Mendelian randomization and implications for interpreting effects on obesity

**DOI:** 10.1101/2023.05.30.23290752

**Authors:** Benjamin Woolf, Héléne T. Cronjé, Loukas Zagkos, Susanna C. Larsson, Dipender Gill, Steve Burgess

**Affiliations:** School of Psychological Science, University of Bristol, Bristol, UK; MRC Integrative Epidemiology Unit, University of Bristol, Bristol, UK; MRC Biostatistics Unit at the University of Cambridge, Cambridge, UK; Department of Public Health, Section of Epidemiology, University of Copenhagen, Copenhagen, Denmark; Department of Epidemiology and Biostatistics, School of Public Health, Imperial College London, London, United Kingdom; Unit of Medical Epidemiology, Department of Surgical Sciences, Uppsala University, Uppsala, Sweden; Unit of Cardiovascular and Nutritional Epidemiology, Institute of Environmental Medicine, Karolinska Institutet, Stockholm, Sweden

## Abstract

Drug-target Mendelian randomization (MR) is a popular approach for exploring the effects of pharmacological targets. *Cis*-MR designs select variants within the gene region that code for a protein of interest to mimic pharmacological perturbation. An alternative uses variants associated with behavioral proxies of target perturbation, such as drug usage. Both have been employed to investigate the effects of caffeine but have drawn different conclusions. We use the effects of caffeine on body mass index (BMI) as a case study to highlight two potential flaws of the latter strategy in drug-target MR: misidentifying the exposure and using invalid instruments. Some variants associate with caffeine consumption because of their role in caffeine metabolism. Since people with these variants require less caffeine for the same physiological effect, the direction of the caffeine-BMI association is flipped depending on whether estimates are scaled by caffeine consumption or plasma caffeine levels. Other variants seem to associate with caffeine consumption via behavioral pathways. Using multivariable-MR, we demonstrate that caffeine consumption behavior influences BMI independently of plasma caffeine. This implies the existence of behaviorally mediated exclusion restriction violations. Our results support the superiority of *cis*-MR study designs in pharmacoepidemiology over the use of behavioral proxies of drug targets.

## Background

Mendelian randomization (MR) is a popular epidemiological study design (1,2). In analogy with randomized controlled trials, MR leverages the random inheritance of genetic variants at conception to improve robustness to reverse causation and confounding. The implementation of MR using an instrumental variables framework has been facilitated by the availability of genome-wide association study (GWAS) summary statistics (3)(4).

Drug-target MR applies this design to explore effects of perturbating pharmacological targets. One method of implementing drug-target MR is to select variants from within the gene region which codes for an exposure of interest (*cis*-variants), in a so called *cis-*MR design (5). While *cis*-MR has the advantage of being mechanistically plausible, it requires understanding of the underlying biology of the drug target.

Studies like the UK Biobank (UKB) have measured participants’ self-reported exposure to many pharmacological compounds (6), such as prescribed and/or over-the-counter medication use, or vitamin supplementation use. These data enable an alternative drug-target MR approach, in which genome-wide significant variants associated with the target respective behavioral proxy are used instead of *cis*-variants (7). Since these phenotypes are generally cheaper to measure than biomarker or protein expression data, they may allow for larger sample sizes and potentially more powerful analyses (8).

Caffeine consumption behavior (e.g. number of cups of coffee or tea consumed in a typical day) has been a popular exposure for studying the effects of increased exposure to caffeine (9–14). *cis*-MR studies using plasma caffeine levels predicted by variants known to affect the metabolism of caffeine have produced contradictory findings to drug-target MR studies using variants robustly associated with self-reported caffeine consumption. For example, observational studies have found that greater exposure to caffeine is predictive of lower body mass index (BMI) and type 2 diabetes mellitus risk (9,10,15). Likewise, the overall results of randomized controlled trials indicate that caffeine intake may promote weight, BMI, and body fat reduction (16). Drug-target MR studies using genome-wide significant variants for caffeine consumption have failed to replicate the inverse association between caffeine consumption and BMI or risk of type 2 diabetes (9–11). Larsson *et al*., however, found results consistent with the observational and trial literature when using *cis*-MR (17). They selected the lead variant association with plasma caffeine levels from the Cytochrome P450 Family 1 Subfamily A Member 2 (*CYP1A2*) and Aryl Hydrocarbon Receptor (*AHR*) gene regions which are known to impact the metabolism of caffeine.

We will explore how the genetic architecture of caffeine consumption behavior relates to that of plasma caffeine levels. We will use this to highlight two potential limitations of using behavioral proxies for understanding the effect of perturbing a pharmacological target: misidentification of the true exposure, and increased risk of invalid instruments.

## Methods

### Study overview

We aim to highlight two possible flaws of using behavioral proxies of target perturbation in a drug-target MR study: 1) misidentifying the direction of effects by not accounting for the mechanism causing the variant-exposure association. 2) increasing risk of including invalid instruments (Figure 1). In the first part of this article, we reproduce the contrasting effects of caffeine on body mass index using *cis*-MR and caffeine consumption instruments. We then use an independent quasi-experimental method, two-way Fixed effects, to adjudicate between them. In the second part of the article, we explore the mechanism underlying variant-caffeine consumption associations and the extent to which this accounts for the contrasting effects. In the final section we explore if the use of a more behavioral phenotype could induce violations of the Instrumental variable (IV) assumptions.

**Figure 1:**
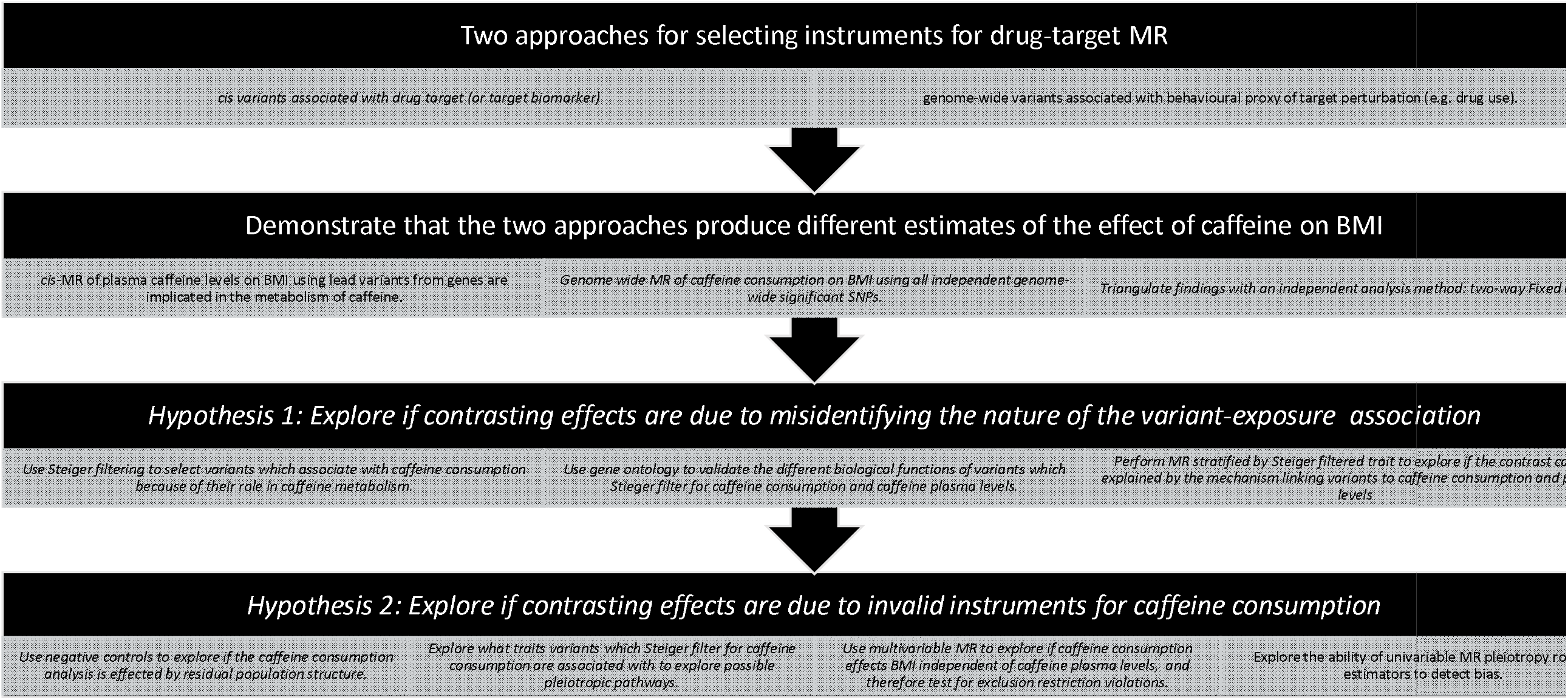
Study overview figure

### Estimating the effects of caffeine on body mass index

#### Genome-wide Mendelian randomization analyses for caffeine consumption

We selected single nucleotide polymorphisms (SNPs) associated with self-reported coffee consumption in the UK Biobank (UKB) as a primary analysis. As a supplementary analysis, we also extracted SNPs associated with self-reported tea consumption in the UKB. These GWASs were conducted according to a standardized pipeline described elsewhere (18). To have sufficient power to detect an effect, we selected independent (clumping r^2^ = 0.001) variants using a 5 × 10^−7^ p-value threshold. Variant-outcome associations were taken from the 2018 GIANT Consortia meta-analysis of BMI, comprising 681,275 participants (19). Variant-trait data were extracted from the OpenGWAS platform using the UKB phenotype IDs: ukb-b-6066, ukb-b-5237, and ieu-b-40 (20). We used the TwoSampleMR R package to harmonise the results, and combined SNP data using an inverse-variance weighted meta-analysis (21).

#### *cis*-Mendelian randomization analysis for plasma caffeine levels

We used the lead variants within the *CYP1A2* and *AHR* gene regions (rs2472297 and rs4410790 respectively). Summary data on the association of these variants with fasting plasma caffeine levels were retrieved from Cornelis *et al*. (22). This was a meta-analysis of 6 studies, collectively including 9,876 European ancestry participants. Variant-outcome data were taken from the same GIANT meta-analysis, and this MR analysis was otherwise conducted using identical methods to the caffeine consumption analyses.

#### Triangulation with two-way fixed effects panel regression

Confidence in a study’s results can be strengthened by triangulating with an alternative design which makes different assumptions (23). One such design is two-way fixed effects (TWFE) panel regression described in Box 1 (24–27).

##### Box 1

Explanation of two-way Fixed Effects

Two-way Fixed Effects (TWFE) is highly popular in the sociology literature (28), and has a similar logic to mono-zygotic twin difference designs used in epidemiology (29). A mono-zygotic twin difference design matches an individual with their mono-zygotic twin who has a discordant exposure status. Given that these twins generally share a developmental environment and possess identical genetics, this matching should remove much confounding bias. Fixed effects (FE) panel regression leverages multiple observations on the same individual at different times to essentially match an individual with themselves at a different time, and therefore control for all time-invariant confounding like genetics. The name derives from the fact that the time-invariant portion of the exposure and outcome is controlled for in the model by estimating it as the fixed effect of the variable’s time series. Time-varying confounding is therefore not adjusted for in a traditional FE analysis. However, when data is available from many time points, it is possible to introduce a fixed effect for time in a TWFE analysis (27). This will adjust for linear time-varying confounding (30,31). More detailed descriptions of FE and TWFE can be found elsewhere (24–28,32,33). Although assuming only linear time-varying confounding is not always plausible, this assumption is very different from the no pleiotropy assumption made by MR, and therefore makes TWFE suitable to triangulate with MR.

We implemented the TWFE model using the fixest R package, and accounted for clustering from both fixed effects in the standard errors (34). Specifically, the UKB research team asked participants to report the number of cups of tea and coffee they drank in a typical day (UKB phenotype IDs: 1488 and 1498) at all four assessment center visits (501,472 participants at recruitment, 20,334 at the first repeat assessment visit, 64,924 at the first imaging visit, and 5,360 at the repeat imaging visit). We standardized the number of cups of tea and coffee measures at each time point and combined them to create a measure of caffeine consumption. One standard deviation of caffeine consumption in the UKB equates to around 2 cups of coffee a day or three cups of tea. At each of these visits, the BMI of each participant was also calculated (UKB phenotype ID: 21001) (6,35).

#### Exploring the genetic architecture of caffeine consumption behavior

One well-measured behavioral phenotype studied using MR is smoking. The genetics of (cigarette) smoking can be split between variants which affect smoking initiation, and those which impact on smoking heaviness (36,37). The Nicotinic receptor genes, for example, inhibit the metabolism of nicotine. While variation in this the gene is not a sensitive predictor of smoking initiation, people who smoke and carry certain variants associated with reduced metabolic inhibition must smoke two to three times more than those without these variants to get the same physiological nicotine effect. Thus, an MR study exploring the effects of nicotine using instruments comprised of variants within genes from a GWAS on smoking heaviness GWAS, could provide counter intuitive estimates, since people who smoke more would have lower genetically predicted nicotine levels.

This vignette closely parallels the explanation provided by Larsson *et al*. for the seemingly discordant effect estimates between their study using plasma caffeine (17), and those using caffeine consumption phenotypes. They found that variants in the *CYP1A2* and AHR genes both predict greater plasma caffeine levels and consumption of less caffeine in the UKB. Since over 95% of UKB participants drank caffeine regularly, one possibility is that is that the instruments associated with higher caffeine consumption are reflective of the need to consume more for caffeine for the same effect due to an increased metabolic efficiency/shorter caffeine half-life. Analogous to the example of nicotine, it may be that people who consume more caffeine do so because they have lower circulating caffeine levels. Since caffeine is not produced endogenously, the opposite conclusion (that caffeine consumption reduces circulating caffeine plasma levels) is biologically implausible.

#### Using Steiger filtering to identify metabolism and behaviorally mediated caffeine consumption SNPs

For the above hypothesis to explain the differences in results we would require most SNPs which associate with caffeine consumption behavior to do so because of their effect on caffeine metabolism. Steiger filtering is a statistical method that determines which of two traits a genetic variant is more likely to primarily affect. The more distal an outcome is from a cause, the less the cause can explain variance in the outcome. Steiger filtering leverages this principle to determine which phenotype is the proximal effect of a genetic variant by comparing the r^2^ of the variant(s) with the phonotypes (38). In its simplest implementation, a SNP will Steiger filter for a trait over a second when it explains a greater percentage of the variation in the first train than the second. We used Steiger filtering to explore which variants are acting on consumption via caffeine metabolism (analogous to smoking heaviness), and which are primarily acting through other pathways. As a supplementary positive control, we used Steiger filtering to objectively confirm that the *CYP1A2* and AHR genes metabolically affect circulating caffeine levels before affecting downstream caffeine consumption behavior.

#### Validating Steiger filtering using gene ontology

To support the biological validity of the conclusions drawn from the Steiger filtering, we explore if the two sets of SNPs have different ontologies. After mapping each variant to its genomic locus, we used the gene ontology database (39) to compare gene overrepresentation between the plasma levels-associated and behavior-associated variants (FDR-adjusted Fisher’s exact tests, PANTHER 17.0, http://pantherdb.org/).

#### SNP mechanism stratified MR to explore exposure misidentification

If the SNPs which Steiger filtered for caffeine plasma levels associate with caffeine consumption because of their role in caffeine metabolism, then we would expect the contrasting effects to be explained by a negative association between circulating plasma caffeine levels. We therefore use MR to first estimate effects between caffeine plasma levels and caffeine consumption for the SNPs which Steiger filtered for each of the respective traits. We also re-estimate the effect of the caffeine consumption SNP (weighted by both caffeine consumption and caffeine plasma levels) on BMI, again stratifying by which trait the SNPs Steiger filtered for.

### Exploring if caffeine consumption variants are invalid instruments

#### Negative controls for population structure

A potential issue with using behavioral proxies for a pharmacological target is the likelihood of a violation of the IV assumptions. It is well established that psychosocial and behavioral GWASs are more prone to residual confounding from population structure, assortative mating, or dynastic effects than biomedical GWASs (40). Without using within family data, care is needed to ensure that MR estimates are not confounded. One method is negative controls (41). Hair color, for example, is a popular negative control outcome for residual population structure, and applicable to our investigation as hair colors differ among population sub-groups, but is unlikely to be caused by caffeine consumption. We extracted UKB GWAS summary statistics on self-reported natural hair color from OpenGWAS (IDs: ukb-d-1747_1, ukb-d-1747_2, ukb-d-1747_6, ukb-b-1997, ukb-b-4263, ukb-b-5593). We then repeated our caffeine consumption analyses (i.e., using any SNP associated with tea or coffee consumption respectively at p < 5 × 10^−7^), but using hair color as the outcome trait.

#### Exploring pleiotropic pathways in behaviorally mediated caffeine consumption SNPs

SNPs acting via behavioral intermediaries are also at greater risk of violating the exclusion restriction assumption. Figure 2 presents two Directed Acyclic Graphs (DAGs) showing plausible mechanisms by which this assumption may be violated for a drug-target MR analysis. In Figure 2a, the variants associate with caffeine consumption through an underlying latent trait which causes both consumption and the outcome, e.g., people that weigh less might consume fewer caffeinated drinks because they lead a healthier lifestyle. In Figure 2b, because caffeinated drinks often contain more than just caffeine there may be an effect of these other substances (e.g., milk, sugar, or an accompanying snack) on BMI, even if we are correctly instrumenting caffeine consumption.

**Figure 2:**
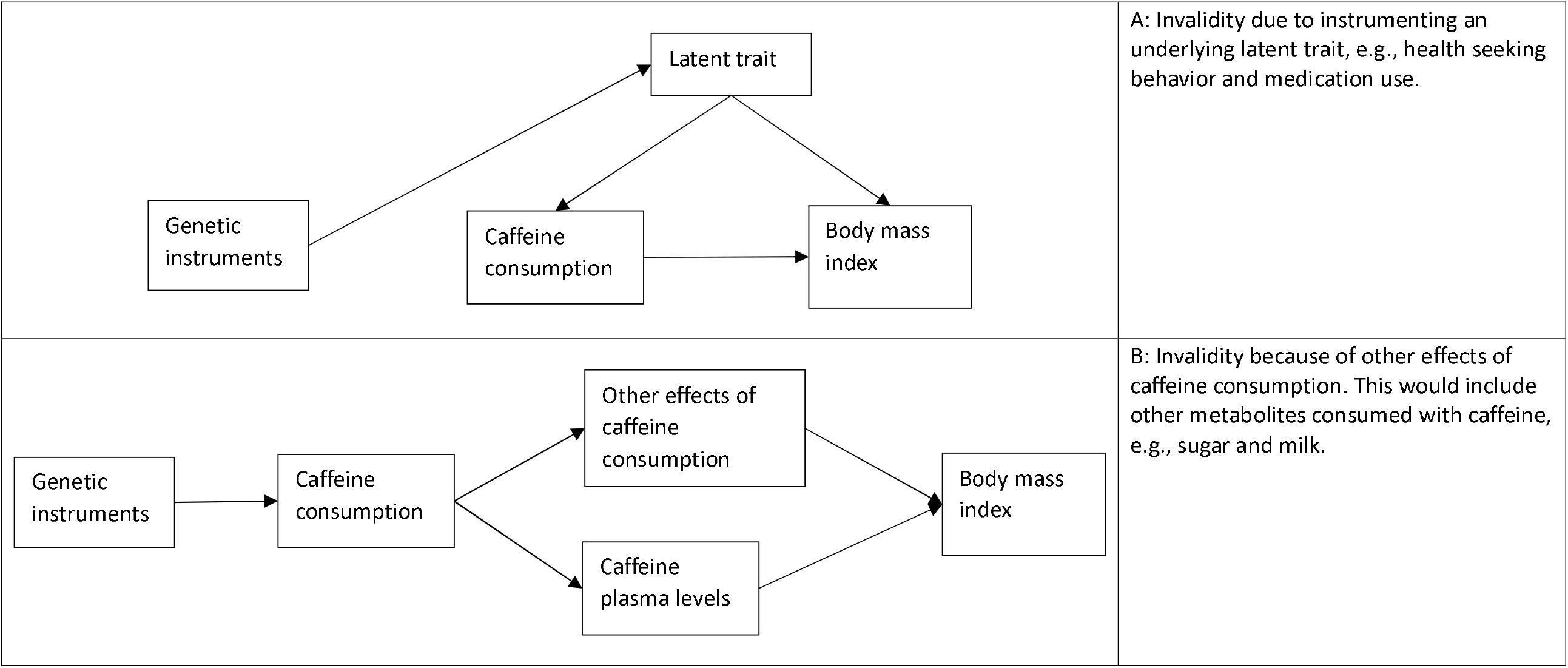
Directed acyclic graphs (DAGs) of potential exclusion restriction violations when using caffeine consumption as an exposure phenotype.

The applicability of DAGs such as those shown in Figure 2 here can be supported by showing that the SNPs which Steiger filter for caffeine consumption are associated with relevant behavioral or lifestyle traits. We therefore use PhenoScanner to explore what traits with these SNPs are associated with (at p < 5 × 10^−5^ FDR, p < 0.05) (42).

#### Multivariable MR to test for exclusion restriction violations in the caffeine consumption MR

If the genetic instruments for caffeine consumption act entirely through the circulating bioactive caffeine metabolite, then there should be no direct effect of caffeine consumption on BMI independent of plasma caffeine levels. Conversely, finding an effect of caffeine consumption independent of caffeine plasma levels would support the existence of exclusion restriction violating pathways like those depicted in Figure 2. We use multivariable MR (MVMR) to test if there is an effect of the caffeine consumption instruments independent of caffeine plasma levels.

MVMR estimates the direct effect of one exposure conditional on another (43). Since we have access to GWAS data on caffeine plasma levels, we can empirically test if caffeine consumption traits act on BMI only through caffeine plasma levels. To perform this analysis, we selected SNPs associated (p < 5 × 10^−7^) with the caffeine consumption or circulating caffeine levels. We then ranked these SNPs in order of their p-values and clumped them with an r^2^ of 0.001. To minimize the effects of conditional weak instrument bias, we implemented the analysis using the MVMR-Qhet estimator, which is more robust to weak instrument bias than inverse variance weighting (47).

#### Univariable pleiotropy robust estimators

Both biases depicted in Figure 2 could be described as pleiotropy, but many pleiotropy robust methods, like MR Egger or weighted median, are likely to produce incorrect estimates in this context: Biases like those in Figure 2a will violate the MR-Egger InSIDE assumption (44), while those in Figure 2b are likely to result in a homogeneous bias across most SNPs which would bias most estimators (45,46). As a final analysis, we explore the ability of commonly used pleiotropy robust estimators (MR-Egger, weighted median, weighted mode, and simple mode (4)) to detect pleiotropy among a) all SNPs associated with coffee consumption, and b) only SNPs which Steiger filter for caffeine consumption.

## Results

### Contrasting Mendelian randomization estimates for the effect of caffeine on BMI

In our MR analysis using self-reported caffeine consumption to construct the instrument, each standard deviation (SD) increase in the genetically-predicted amount of coffee consumed was associated with 0.754 (95% CI: 0.284 to 1.224) SD higher BMI. This analysis had variant-exposure F statistic of 239.

Our *cis*-MR study (instrumenting on plasma caffeine levels) had a variant-exposure F statistic of 71. We found that each SD increase in genetically-predicted plasma caffeine levels associated with a 0.085 SD decrease (95% CI: -0.095 to -0.075) in BMI.

The TWFE regression model found that each SD increase in caffeine consumption associated with a 0.021 (95% CI: -0.032 to -0.010) kg/m^2^ *reduction* in BMI. This therefore supports the conclusion of the *cis*-MR analysis.

### Implications of the genetic architecture of caffeine consumption on Mendelian randomization estimates

Steiger filtering on the variants associated with caffeine consumption behavior implied that 16 of the 24 coffee-consumption-associated SNPs are affecting caffeine consumption behavior because of their effect on caffeine metabolism. This interpretation was supported by the gene ontology analysis. We found that plasma caffeine level-associated variants were significantly enriched for the biological process response to *organic substance* (3.43-fold enrichment, p = 8.67 × 10^−7^). However, no statistically significant gene enrichment was observed for consumption-associated variants. This supports there being a less directly biological (e.g. a behavioral) mechanism linking these other variants to caffeine consumption.

The SNPs which Steiger filtered for caffeine plasma levels imply a negative MR association between circulating plasma caffeine levels and caffeine consumption (beta = -0.295 SD consumed per SD increase in plasma caffeine levels, 95% CI: -0.403 to -0.187), replicating the observation by Larsson *et al*. Thus, although using these SNPs to estimate the effect of caffeine consumption on BMI produces a positive MR estimate (beta = 0.586 SD per SD increase in coffee consumption, 95% CI: 0.047 to 1.125), subsequent scaling by plasma caffeine levels results in similar MR estimates (beta = -0.141 SD per SD increase in plasma caffeine levels, 95% CI: -0.353 to 0.071) to those observed in the *cis*-MR analysis above.

The remaining SNPs are more proximal to caffeine consumption behaviour than plasma caffeine levels. The MR analysis of caffeine consumption on BMI using these SNPs still implies that increased caffeine consumption may increase BMI (beta = 1.541 SD per SD increase in coffee consumption, 95% CI: 0.610 to 2.472). This cannot be explained by the effect these SNPs have on circulating caffeine: MR of caffeine consumption on plasma caffeine levels are indicative of a positive direction of effect (beta = 0.416 SD per SD increase in coffee consumption, 95% CI: 0.110 to 0.722).

### Caffeine consumption variants can be invalid instruments

We first used hair color as a negative control outcome for population structure when using the caffeine consumption instruments. This analysis failed to find evidence of an association of the instruments with these traits (Supplementary Table S1), so it seems unlikely that the caffeine consumption GWASs are biased by residual population structure.

However, many of the SNPs which Steiger filtered for caffeine consumption are associated (p < 5 × 10^− 5^) with behavioral traits such as smoking, alcohol consumption, education, and physical activity (and BMI related traits), represented in PhenoScanner (Supplementary Tables S2a and S2b). Any of these could be a source of exclusion restriction violations.

Indeed, in our MVMR model, we find some evidence of a direct effect of coffee consumption, independent of plasma caffeine levels, on BMI. Each SD increase in coffee consumption results in a 0.556 (95% CI: 0.296 to 1.829) SD increase in BMI, independent of plasma caffeine levels.

Despite MVMR implying the existence of meaningful exclusion restriction violations, the traditional pleiotropy robust methods still generally imply a positive causal effect of caffeine consumption on BMI when using SNPs Steiger filtered for caffeine drinking behavior (Supplementary Table S3a and S3b).

Results of the supplementary analysis using data from the UKB GWAS of tea consumption are similar to those using the GWAS of coffee consumption and can be found in the supplementary results.

## Discussion

In this paper we explored, and found evidence to support, two hypotheses for the differences observed between *cis*-MR estimates for the effect of caffeine plasma levels on BMI, and drug-target MR estimates for the effect of caffeine consumption on BMI. Specifically, we explore if the caffeine consumption variants are misidentifying the true exposure, and if instruments selected by these GWASs are in fact invalid.

To explore the first hypothesis, we performed Steiger filtering to identify which SNPs associate primarily with caffeine plasma levels or caffeine consumption. Around half the SNPs for caffeine consumption appear to affect caffeine consumption because of their effect on caffeine metabolism. Counterintuitively, we observed a negative MR association between genetically predicted caffeine plasma levels and caffeine consumption. Since caffeine consumption cannot cause lower caffeine plasma levels, this is probably due to people with elevated genetically predicted plasma caffeine needing to drink less coffee or tea. Indeed, gene enrichment analysis revealed overrepresentation of genes involved in the metabolic response to the presence of organic substances, specifically processes resulting in physiological tolerance to an organic substance, among these SNPs. When scaling the MR effect using consumption-related SNPs on BMI by their concurrent effect on caffeine plasma levels, we find a similar association to that in the *cis*-MR analysis. This means that a naive interpretation of MR using caffeine consumption to proxy caffeine plasma levels may produce misleading results and demonstrates the importance of understanding the biological mechanism linking a behavioral phenotype to the drug-target biomarker.

The MR analyses of caffeine consumption on both BMI and plasma caffeine using SNPs which Steiger filtered for consumption behavior find that increased caffeine consumption increases BMI. This cannot be due to the misidentification of caffeine metabolism variants as caffeine consumption variants because the direction of effect is identical in both MR analyses. Instead, we argue that these remaining SNPs are likely to produce invalid MR estimates due to violations of the exclusion restriction assumption. This hypothesis is supported by the range of behavioral phenotypes these traits are associated with in PhenoScanner. Indeed, we were able to demonstrate the existence of a causal effect on BMI independent of caffeine plasma levels using MVMR. This implies that a drug-target MR using these variants would suffer from an exclusion restriction violation. Thus, SNP validity, in addition to specification of the correct exposure, can complicate the interpretation of the caffeine consumption MR effect estimates.

Finally, we triangulated with two-way fixed effect panel regression to test whether *cis*-MR estimates are reliable. Since the assumption of no non-linear time varying confounding needed by TWFE is very different to the MR assumption of no pleiotropy, this result should lend extra credence to the finding that caffeine consumption results in weight loss.

The failure of commonly used pleiotropy robust methods to produce estimates with the correct direction of effect demonstrates the difficulty in detecting systematic pleiotropy in settings with complex behavioral exposures. These estimators typically assume that each SNP has an idiosyncratic pleiotropic effect. Researchers therefore need to be cautious not to overinterpret findings in settings where similar exclusion restriction violations may affect a large proportion of SNPs. Positive and negative controls should be used to detect if related traits could result in violations of the exclusion restriction assumption. For example, Wang *et al*. incorporated supplementation use known to not effect diabetes as a negative control exposure for an effect of generic supplementation use in a MR study exploring the effect of zinc supplementation on diabetes risk (7). Examples for caffeine consumption could be UKB GWASs of green tea consumption, as green tea is typically consumed without milk and sugar. Alternatively, the consumption of decaffeinated tea or coffee could be investigated when attempting to separate the caffeine-specific effect from the effect of typically added substances like milk and sugar. Currently, however, available GWASs of these traits are not adequately powered to be useful as negative control exposures (48–50).

Our study has assumed that the research question of interest is the effects of caffeine itself, rather than caffeine consumption. A recent MR study found an association between genetically proxied coffee consumption and esophageal cancer (13). This result was interpreted as being because of consuming hot liquids, rather than caffeine. While Figure 2b is a suitable description of this interpretation of this effect it should not be described as resulting from an exclusion restriction violation. In such a study, the behavior, rather than the drug target, is the exposure of interest. As such, the relative merits of different study designs depend on the research question being answered. When the mechanism linking metabolism and consumption behavior is understood, there may still be utility in using *cis* designs to explore the effect of consumption behavior because they might avoid biases like those depicted in Figure 2a. Indeed, one interpretation of the difference between the direction of the MR-Egger estimates in Supplementary Table S3 and our TWFE estimates is that the MR-Egger InSIDE assumption has been violated. The consistency between the genome-wide and *cis* MR results in the aforementioned oesophageal cancer MR analysis, on the other hand, imply that this may not be a major threat to this study’s validity.

The existing literature has already noted some issues, such as confounding by indication, when using behavioral exposures to proxy pharmacological interventions in an drug-target MR framework (51). Likewise, the gene-environment equivalence assumption, which is required for MR estimates to translate to the effects of pharmacological interventions, is less plausible when using these behavioral phenotypes (52,53). We believe that the two issues we have highlighted here could be relevant for other pharmacological targets. Since the relationship between caffeine consumption and plasma caffeine levels should be relatively simple – caffeine is drunk by nearly all participants in the UKB and is not created endogenously – our results demonstrate that caution is required when interpreting drug-target MR studies using a behavioral exposure to proxy a pharmacological target. While careful thought is always required when choosing instruments and exposure traits for drug-target MR studies (54), we believe that our results support the superiority of *cis*-MR study designs in pharmacoepidemiology over the use of behavioral proxies of drug targets.

**Table 1:** Summary and interpretation of the study’s results

| Analysis | Exposure (units) | Outcome (units) | Variant-exposure F statistic | Number of SNPs | Effect (95% CI) | Interpretation |
| --- | --- | --- | --- | --- | --- | --- |
| Genome-wide MR | Coffee consumption (SD) | BMI (SD) | 239 | 24 | 0.754 (0.284 to 1.224) | Greater caffeine consumption associates with weight gain using genome-wide MR. |
| *cis*-MR | Plasma caffeine levels (SD) | BMI (SD) | 71 | 2 | -0.085 (-0.095 to -0.075) | Exposure to more caffeine results in weight loss. Since TWFE and *cis*-MR make very different assumptions, the triangulation between the two estimates supports the validity of the *cis*-MR analysis. |
| two-way Fixed effects (TWFE) | Caffeine consumption (SD) | BMI (SD) | NA | NA | -0.100 (-0.152 to -0.048) |  |
| Genome-wide MR using SNPs which Steiger filter for caffeine plasma levels | Plasma caffeine levels (SD) | Coffee consumption (SD) | 11 | 16 | -0.295 (-0.403 to -0.187) | Caffeine consumption SNPs which Steiger filtered for caffeine plasma levels imply a positive effect of caffeine consumption on BMI. However, when scaled by plasma caffeine levels they produce similar estimates to the *cis*-MR. Since caffeine consumption is unlikely to reduce caffeine plasma levels, which the MR implies, it is likely that these SNPs measure the effect that caffeine metabolism has on caffeine consumption. |
|  | Coffee consumption (SD) | BMI (SD) | 112 | 16 | 0.586 (0.047 to 1.125) |  |
|  | Plasma caffeine levels (SD) | BMI (SD) | 112 | 16 | -0.141 (-0.353 to 0.071) |  |
| Genome-wide MR using only SNPs which Steiger filter for caffeine consumption | Coffee consumption (SD) | BMI (SD) | 42 | 8 | 1.541 (0.610 to 2.472) | Caffeine consumption SNPs which Steiger filtered for caffeine consumption imply a positive effect of caffeine consumption on BMI. Since they also have a positive effect on caffeine plasma levels, this cannot be explained by miss-identification of the exposure. However, these SNPs do associate with behavioral phenotypes which could also effect BMI (Supplementary Table 2) |
|  | Coffee consumption (SD) | Plasma caffeine levels (SD) | 42 | 8 | 0.416 (0.110 to 0.722) |  |
| Multivariable MR (using the MVMR-Qhet estimator) | Direct effect of caffeine consumption (SD) after adjusting for plasma caffeine levels | BMI (SD) | 5 (plasma caffeine) and 42 (caffeine consumption) | 25 | 0.556 (0.296 to 1.829) | Caffeine consumption has a direct effect on BMI, independent of caffeine plasma levels. This implies that drug target MR using caffeine consumption to study the effects of caffeine could suffer from exclusion restriction violations |
| Genome-wide MR using pleiotropy robust | Coffee consumption (SD) | BMI (SD) | 239 | 24 | Generally positive (Supplementary | Over interpretation of pleiotropy robust estimators may be misleading in the presence of systematic exclusion restriction violations |
| estimators |  |  |  |  | Table 3) |  |

## Data Availability

All data produced are available online.

## Author contributions

B.W. and T.C. performed statistical analysis. B.W. wrote the first draft of the manuscript. All authors contributed to the design of the study and edited the manuscript for intellectual content.

## Funding

B.W. is funded by an Economic and Social Research Council (ESRC) South West Doctoral Training Partnership (SWDTP) 1 + 3 PhD Studentship Award (ES/P000630/1) and a is member of the Medical Research Council (MRC) Integrative Epidemiology Unit at the University of Bristol (MC_UU_00011/7). D.G. is supported by the British Heart Foundation Centre of Research Excellence (RE/18/4/34215) at Imperial College. S.C.L. is supported by the Swedish Research Council for Health, Working Life and Welfare (Forte, 2018-00123), Swedish Heart Lung Foundation (Hjärt-Lungfonden, 20210351), and Swedish Research Council (Vetenskapsrådet, 2019-00977).

## Ethics Approval statement

UKB received ethics approval from the North West Multi-Centre Research Ethics Committee (REC reference 11/NW/0382). All participants provided written informed consent to participate in the study. Data from the UKB are fully anonymized.

## Declaration of interest

DG is employed part-time by Novo Nordisk. The other authors declare no conflicts of interest.

## Acknowledgments

*This work was carried out using the computational facilities of the Advanced Computing Research Centre, University of Bristol -http://www.bris.ac.uk/acrc/*.

This project was conducted using UK Biobank application no. 15825. UK Biobank was established by the Wellcome Trust medical charity, Medical Research Council, Department of Health, Scottish Government and the Northwest Regional Development Agency. It has also had funding from the Welsh Government, British Heart Foundation, Cancer Research UK and Diabetes UK. UK Biobank is supported by the National Health Service (NHS). UK Biobank is open to bona fide researchers anywhere in the world.

## Supplementary Results

### Replication of coffee consumption results using tea consumption

We were able to broadly replicate the coffee consumption findings in the main text using the UKB GWAS of tea consumption. We found, in our MR using self-reported tea consumption, that each standard deviation (SD) increase in the genetically-predicted amount of tea consumed was associated with 0.225 (95% CI: 0.045 to 0.405) SD higher BMI. This analysis had variant-exposure F statistic of 104.

Steiger filtering on the variants associated with caffeine consumption behavior implied that 10 of the 26 tea consumption-associated SNPs are affecting caffeine consumption behavior because of their effect on caffeine metabolism. As with the coffee analysis, the SNPs which Steiger filtered for caffeine plasma levels imply a negative MR association between circulating plasma caffeine levels and tea consumption (beta = -0.320, 95% CI = -0.465 to 0.175). Again, when using these SNPs to estimate the effect of tea consumption on BMI produces a positive MR estimate (beta = 0.256, 95% CI = 0.146 to 0.366), but scaling by plasma caffeine levels result in similar MR estimates (beta = -0.064, 95% CI = -0.131 to 0.003) to those observed in the cis-MR analysis above.

The MR analysis of tea consumption on BMI using SNPs which Steiger filter for tea consumption still implies that increased tea consumption may increase BMI (beta = 0.150, 95% CI = -0.254 to 0.554). This again cannot be explained by the effect these SNPs have on circulating caffeine: MR of tea consumption on plasma caffeine levels are indicative of a positive direction of effect (beta = 0.026, 95% CI = -0.239 to 0.291).

Our MVMR model again finds some evidence of a direct effect of tea consumption, independent of plasma caffeine levels, on BMI: Each SD increase in tea consumption results in a a 0.280 (95% CI: 0.073 to 0.568) SD increase in BMI independent of plasma caffeine levels. The conditional F statistics for caffeine plasma levels and tea consumption were 4 and 8 respectively.

### Positive control analysis for Steiger filtering

Our positive control analysis for Steiger filtering confirmed that *the CYP1A2* and AHR genes influence circulating caffeine levels (r2 = 1.4% for both genes combined) before either caffeine consumption behavior (r2 = 0.2% for both genes combined).

## Notes

### Author Declarations

OpenGWAS project or GWAS original publications.

